# Medical Students’ Use of Large Language Models: A National Survey

**DOI:** 10.64898/2026.01.26.26344898

**Authors:** Austin A. Barr, Robert C. Rozman, Kevin Liu, Michelle Pham, Zachary Klarenbach, Arun Chinna-Meyyappan, Ahmad Y. Hassan, Michael Zarychta, Omar El Ferri, Ali Al-Khaz’Aly, Praneesh Datt, Aydin I. Herik, Kareem Sadek, Mike Paget, Jessalyn K. Holodinsky

## Abstract

**Background:** Large language models (LLMs) are increasingly embedded in medical education and clinical care settings, yet limited empirical data describe medical students in Canada’s use and perceptions of these tools. We aimed to characterize student engagement including LLMs used, frequency, purposes, trust, accuracy, perceived impacts, and attitudes toward educational and clinical integration.

**Methods:** We conducted a national survey of medical students in Canada distributed between November and December 2025. We summarized responses using descriptive statistics and compared results between students in preclerkship versus clerkship using Fisher’s exact test.

**Results:** Among 286 respondents from 10 medical schools, 96.50% reported using at least one LLM. The most commonly used LLMs were ChatGPT (93.36%) and OpenEvidence (57.69%). Daily/weekly use was most frequent for coursework assistance (60.22%) and clinical questions (57.14%). Most respondents reported positive impacts on efficiency (81.62%), learning (77.01%), and academic performance (59.49%). Students commonly reported encountering inaccurate information (90.18%). Formal instruction on LLM use was uncommon (10.95%), though 67.67% of students agreed medical schools should integrate formal instruction on LLMs. Only 21.43% of respondents felt adequately educated on data privacy regulations applicable to these tools.

**Conclusions:** LLM use among surveyed medical students in Canada was nearly universal and perceived favourably. However, students reported exposure to inaccurate outputs and substantial gaps in formal training and privacy literacy. These findings support the development of structured curricular guidance on appropriate application of these tools, including information verification practices and ethical, privacy-aware engagement.

## Introduction

Large language models (LLMs), a widely accessible form of generative artificial intelligence (AI), can generate fluent, context-relevant text in response to natural-language prompts. In medicine, LLMs are increasingly used to synthesize information, draft written materials, and support conversational question answering [1,2,3].

Early work evaluating LLM performance on medical knowledge tasks accelerated attention to these tools within health professions training. In particular, studies which evaluated general-purpose LLMs on United States Medical Licensing Examination (USMLE)-style questions prompted discussion on utility toward medical education [4]. More recently, domain-adapted medical LLMs have reported even higher performance on USMLE-style benchmarks and subspecialty clinical vignettes [5].

Beyond knowledge assessment, published work increasingly reflects experimentation with LLMs to support learning experiences, academic workflows, and research. Interactive LLM-based tools have been developed to support clinical skills practice through simulated patient encounters and objective structured clinical examination (OSCE)-oriented learning [6]. In parallel, LLMs have been evaluated for generating medical examinations and question banks [7,8], and to support learners through clinical question-answering settings [9]. LLMs are also being explored to accelerate components of evidence synthesis [10], generate clinical data [11,12], facilitate clinical documentation [13], and to support clinical reasoning [14]. Together, the literature suggests that LLMs are evolving into a broad ecosystem of tools that can influence how students learn, how educators assess, and how future clinicians interact with information and documentation.

Despite promising applications, medical educators and clinical training programs are simultaneously navigating questions about acceptable use, transparency, and risk. Core concerns include inaccurate outputs, ‘hallucinations’, bias, variability in performance across prompts and contexts, academic integrity challenges, and the long-term impacts on student learning [1,15-17]. Privacy and security risks are also salient, particularly if users input sensitive information into third-party systems with unclear data retention or secondary use policies [18,19]. In clinical learning contexts, these issues intersect with professionalism and confidentiality norms, creating uncertainty about what constitutes appropriate use.

A current, comprehensive understanding of how medical students engage with LLMs remains limited, leaving educators without the empirical basis to set clear expectations, manage risks, and guide appropriate integration into medical training. Surveys in other jurisdictions document rapid adoption among medical learners [20,21], yet Canadian data lags. To address this gap, we conducted a national survey study of English-speaking medical students in Canada with the objectives of exploring the prevalence and frequency of LLM use, identifying which LLMs are used and for what purposes, assessing student perceptions regarding utility, accuracy, limitations, and impact, and to describe perceived barriers, challenges, and ethical concerns.

## Methods

We conducted a cross-sectional, anonymous survey of medical students in Canada between November and December 2025. Eligibility included enrollment in a Canadian MD program and current stage of training categorized as preclerkship or clerkship.

### Survey Instrument Development

The survey instrument was developed, in English, through an iterative process informed by a review of the existing literature on LLM use in medical education and higher education, as well as consultation with medical trainees to ensure relevance and clarity. Initial items were drafted to capture patterns of LLM access and use, perceived benefits and risks, and attitudes toward integration in educational and clinical settings. The drafted survey was pilot tested with a small group of medical students from different training stages to assess question clarity, content validity, and completion time. Feedback from pilot testing was used to refine item wording, response options, and survey structure prior to national dissemination. No personally identifying information was collected during pilot testing.

### Recruitment and Data Collection

The survey was disseminated via official medical school channels, medical student unions, and national/regional medical student groups (i.e., Canadian Federation of Medical Students and Canadian Atlantic Medical Students Association) to English-speaking medical students. The survey was administered using the Qualtrics survey platform (Qualtrics, Provo, UT). As an incentive, participants could enter a random prize draw for one of ten $20 Amazon gift cards.

### Survey Instrument and Measures

The survey instrument assessed: (1) respondent characteristics (training stage and institution); (2) LLM exposure and access (models used, paid subscription status, whether LLM use had ever been required during medical training, and whether they encountered LLM-generated content during schooling); (3) frequency of LLM use for specific tasks (coursework assistance, writing assistance, clinical questions, research, written exam preparation, and OSCE preparation); (4) perceived impacts on learning, academic performance, and efficiency; (5) perceptions of trust and accuracy, and frequency of encountering inaccuracies; and (6) agreement with statements regarding curriculum integration, access barriers, data privacy preparedness, and ethical concerns in educational and clinical contexts. The full survey instrument is provided in the Supplement (Supplementary Appendix 1).

### Statistical Analysis

Descriptive statistics were calculated for recorded variables. Categorical data were summarized as counts and percentages. Statistical associations between categorical variables were evaluated using Fisher’s exact test. Statistical analyses were performed in R (version 4.4.2, The R Project for Statistical Computing), and figures were created using the Python library Matplotlib (https://matplotlib.org).

### Ethics

Ethics approval was obtained from the Conjoint Health Research Ethics Board (CHREB), University of Calgary, with additional institutional approvals obtained where required.

## Results

### Respondent characteristics

A total of 286 English-speaking medical students responded, with participants representing 10/18 of Canada’s medical schools. Most respondents were in the preclerkship (235/286; 82.17%) stage of medical training. The majority of respondents were enrolled at the University of British Columbia (99/286; 34.62%), University of Calgary (81/286; 28.32%), and the University of Manitoba (36/286; 12.59%). The remaining 70 students (24.48%) were distributed amongst the University of Ottawa, University of Alberta, University of Toronto, Memorial University of Newfoundland, Western University, Dalhousie University, and Northern Ontario School of Medicine.

### LLM Adoption and Models Used

Overall, 276/286 (96.50%) students reported using at least one LLM (Table 1). The most commonly used models were ChatGPT (267/286; 93.36%) and OpenEvidence (165/286; 57.69%). OpenEvidence use was more frequent among students in clerkship than preclerkship (70.59% vs 54.89%). Less than one third of respondents (81/273; 29.7%) reported having a paid subscription to an LLM, but if so the most commonly reported was ChatGPT.

**Table 1.** Large language models used, overall and by training stage (multi-select; N=286)

| **Large Language Model** | **Overall**<br>**n (%)** | **Preclerkship**<br>**n (%)** | **Clerkship**<br>**n (%)** |
| --- | --- | --- | --- |
| ChatGPT | 267 (93.36) | 220 (93.62) | 47 (92.16) |
| OpenEvidence | 165 (57.69) | 129 (54.89) | 36 (70.59) |
| Gemini | 72 (25.17) | 63 (26.81) | 9 (17.65) |
| CoPilot | 29 (10.14) | 26 (11.06) | 3 (5.88) |
| DeepSeek | 29 (10.14) | 23 (9.79) | 6 (11.76) |
| Claude | 15 (5.24) | 12 (5.11) | 3 (5.88) |
| Perplexity | 11 (3.85) | 8 (3.40) | 3 (5.88) |
| Grok | 10 (3.50) | 9 (3.83) | 1 (1.96) |
| NotebookLM | 10 (3.50) | 9 (3.83) | 1 (1.96) |
| Mistral | 3 (1.05) | 3 (1.28) | 0 (0.0) |
| Llama | 2 (0.70) | 2 (0.85) | 0 (0.0) |
| Consensus | 1 (0.35) | 1 (0.43) | 0 (0.0) |
| None | 10 (3.50) | 7 (2.98) | 3 (5.88) |

### Educational Exposure and Training

Seventeen percent of respondents (50/286; 17.48%) reported having been required to use an LLM as part of their medical schooling, with higher prevalence in preclerkship (20.00%) compared with clerkship (5.88%) (*p* = 0.014) (Table 2). Self-reported comfort using LLMs was generally high, but did not differ by training stage (*p* = 0.712). More than half (151/273; 55.31%) of respondents indicated encountering LLM-generated content from instructors, with significant difference by training stage (*p* = <0.001). Only 10.95% (30/274) reported having received formal training or instruction on how to use LLMs, with 4.01% (11/274) reporting training directly from school; however, formal training was not consistently reported at any single surveyed institution.

**Table 2.**
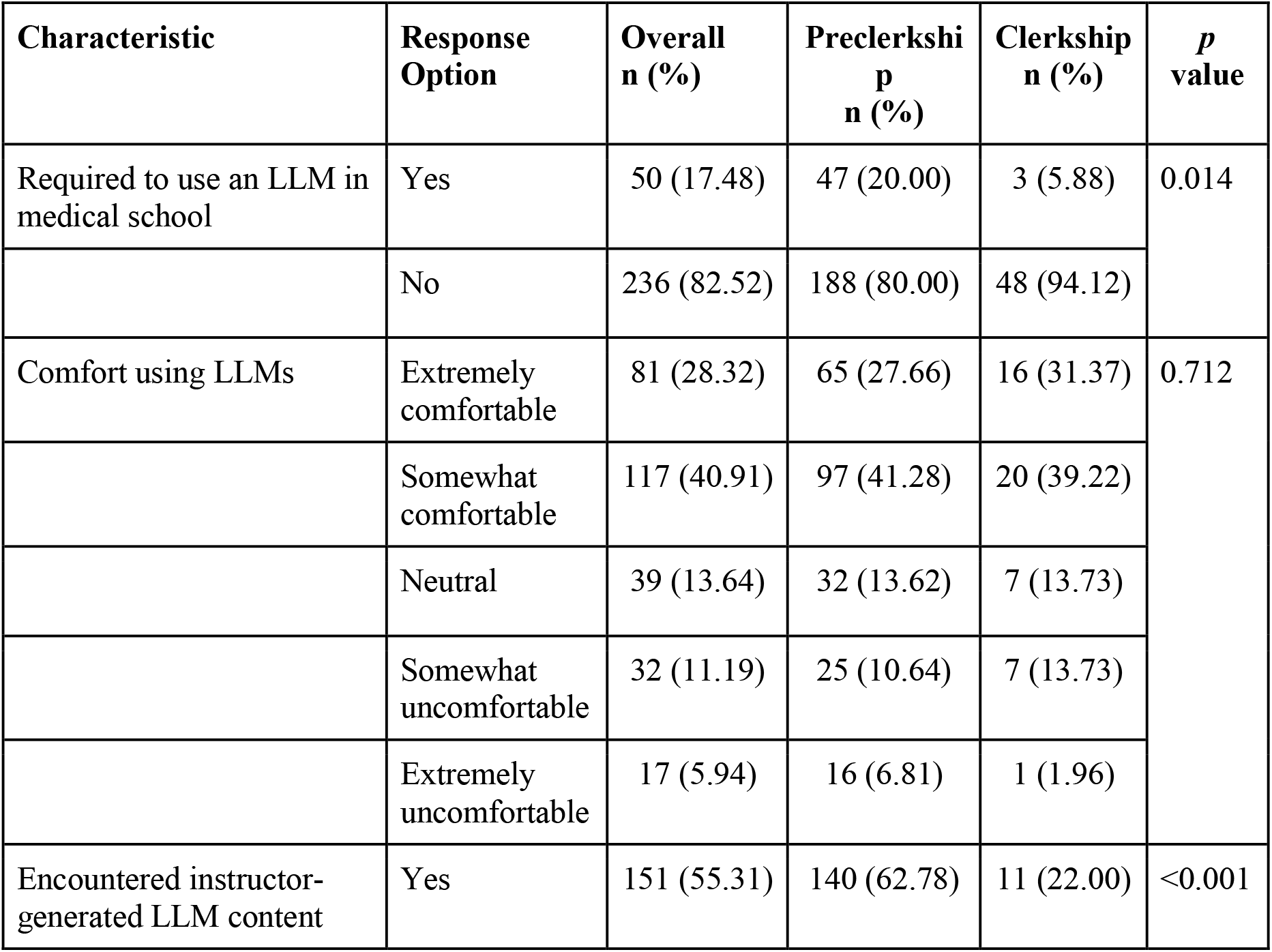
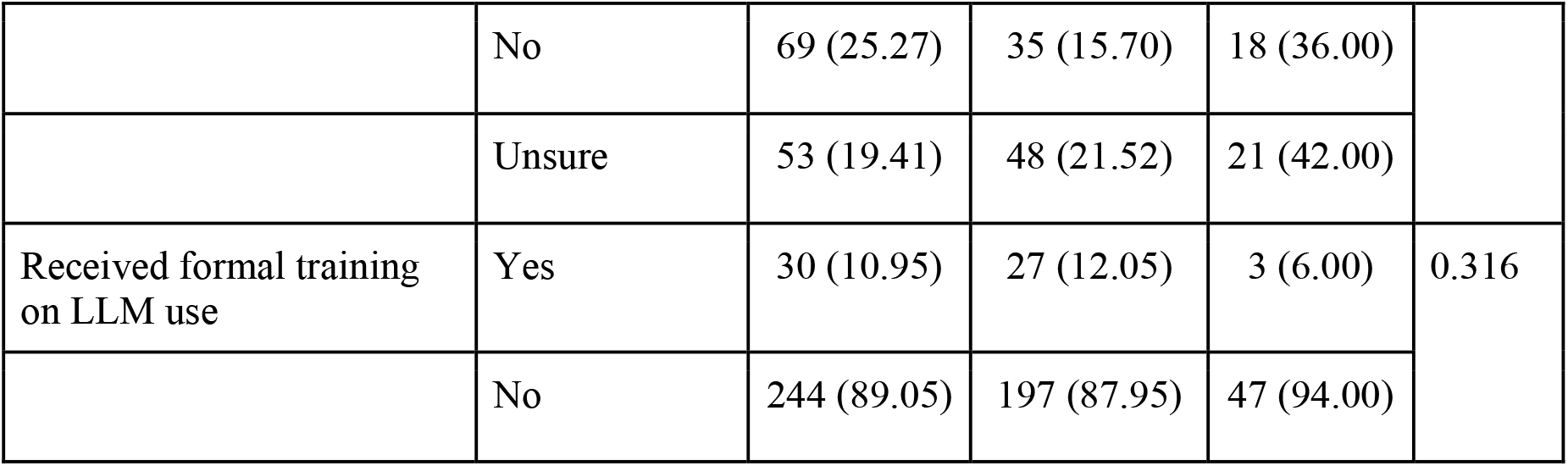
Educational exposure and training related to large language models (LLMs)

### Frequency of LLM use by Task

Across learning activities, daily/weekly use was most common for coursework assistance (165/274; 60.22%) and for answering clinical questions (156/273; 57.14%) (Figure 1). Frequency distributions differed by training stage for coursework assistance (*p* = 0.011) and OSCE preparation (*p* = 0.028), with preclerkship respondents reporting more frequent use.

**Figure 1.**
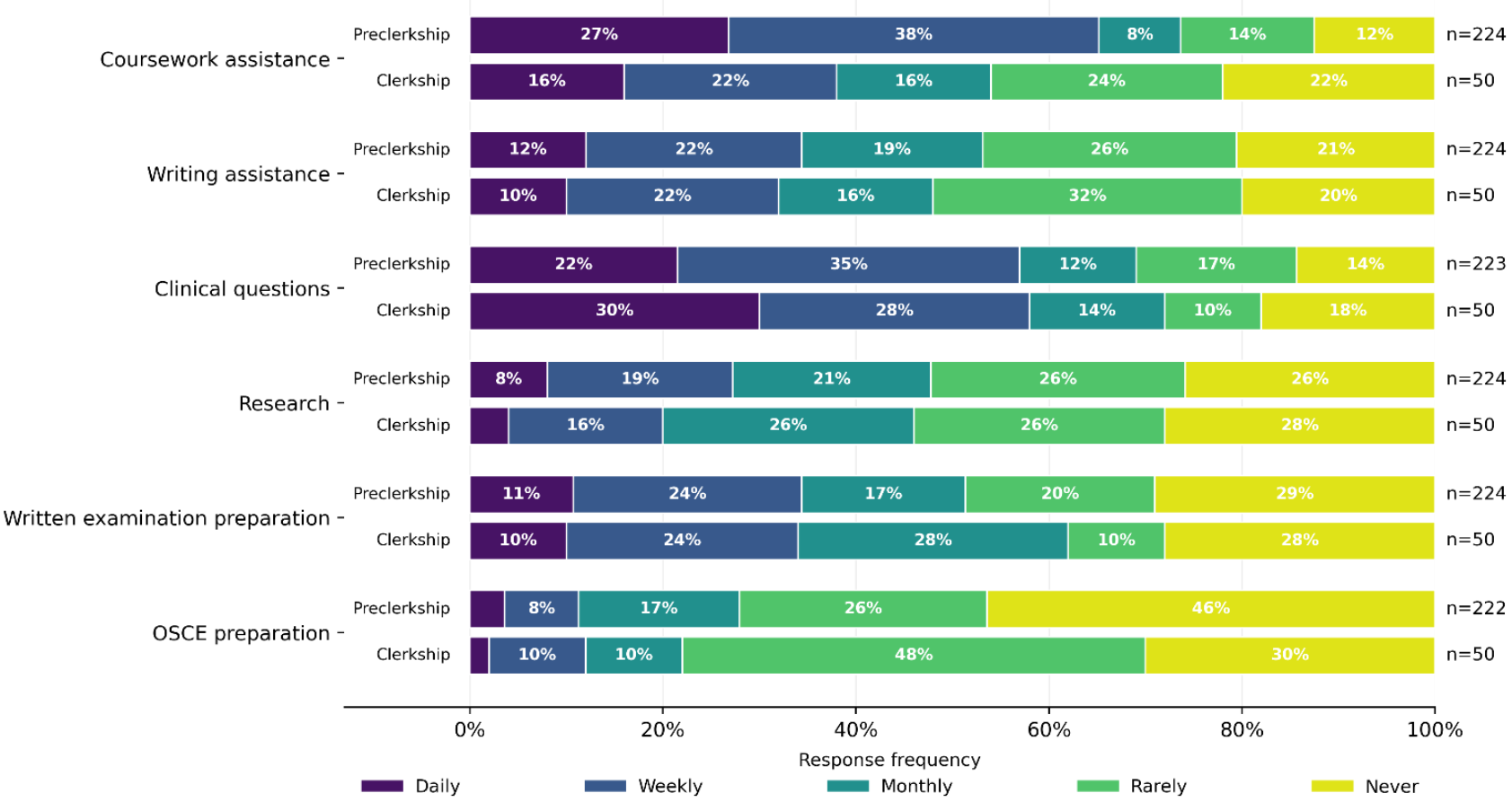
Frequency of large language model use by task, stratified by training stage (preclerkship vs clerkship).

### Perceived Impacts, Trust, and Accuracy Perceptions

Most respondents reported positive impacts on learning (211/274; 77.01%), academic performance (163/274; 59.49%), and efficiency (222/272; 81.62%) (Figure 2). Perceived positive impact on academic performance had a significant difference by training stage (*p* = 0.038), with a higher proportion of preclerkship students reporting positive impact.

**Figure 2.**
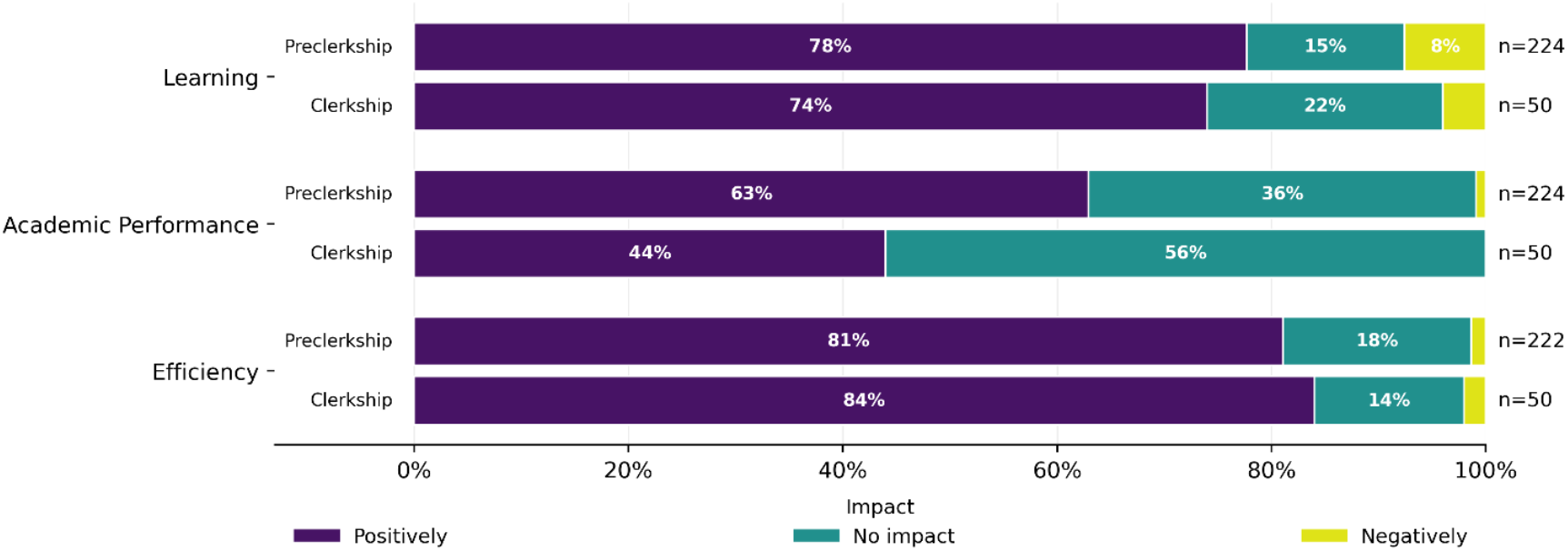
Self-reported impact of large language models on learning, academic performance, and efficiency, stratified by training stage (preclerkship vs clerkship).

When asked to compare LLMs with traditional learning resources, 175/275 (63.64%) of students rated LLMs as somewhat or much better (Table 3). Trust in LLM-provided information was most commonly rated as “mostly” or “somewhat” (205/275; 74.55%), with no respondents indicating “completely.” A large majority of students reported encountering inaccurate information from LLMs (248/275; 90.18%), most commonly reported as occurring “once in a while” (176/262; 67.18%).

**Table 3.** Perceived impacts, trust, and accuracy perceptions.

| Characteristic | Response Option | Overall n (%) | Preclerkship n (%) | Clerkship n (%) | p value |
| --- | --- | --- | --- | --- | --- |
| LLMs compared with traditional learning resources | Much better | 56 (20.36) | 46 (20.44) | 10 (20.00) | 0.056 |
|  | Somewhat better | 119 (43.27) | 92 (40.89) | 27 (54.00) |  |
|  | About the same | 50 (18.18) | 47 (20.89) | 3 (6.00) |  |
|  | Somewhat worse | 37 (13.45) | 28 (12.44) | 9 (18.00) |  |
|  | Much worse | 13 (4.73) | 12 (5.33) | 1 (2.00) |  |
| Trust in LLM-provided information | Completely | 0 (0.0) | 0 (0.0) | 0 (0.0) | 0.152 |
|  | Mostly | 97 (35.27) | 76 (33.78) | 21 (42.00) |  |
|  | Somewhat | 108 (39.27) | 95 (42.22) | 13 (26.00) |  |
|  | A little | 47 (17.09) | 37 (16.44) | 10 (20.00) |  |
|  | Not at all | 23 (8.36) | 17 (7.56) | 6 (12.00) |  |
| Encountered inaccurate information from LLMs | Yes | 248 (90.18) | 200 (88.89) | 48 (96.00) | 0.408 |
|  | No | 6 (2.18) | 6 (2.67) | 0 (0.0) |  |
|  | Unsure | 21 (7.64) | 19 (8.44) | 2 (4.00) |  |
| Frequency of encountering inaccuracies | Every prompt | 1 (0.38) | 1 (0.47) | 0 (0.0) | 0.098 |
|  | Every few prompts | 36 (13.74) | 33 (15.57) | 3 (6.00) |  |
|  | Once in a while | 176 (67.18) | 136 (64.15) | 40 (80.00) |  |
|  | Rarely | 47 (17.94) | 41 (19.34) | 6 (12.00) |  |
|  | Never | 2 (0.76) | 1 (0.47) | 1 (2.00) |  |

Of the LLMs respondents indicated they used, they were asked to rank each in order of trust. OpenEvidence was most frequently rated highest among respondents on trust (Figure 3).

**Figure 3.**
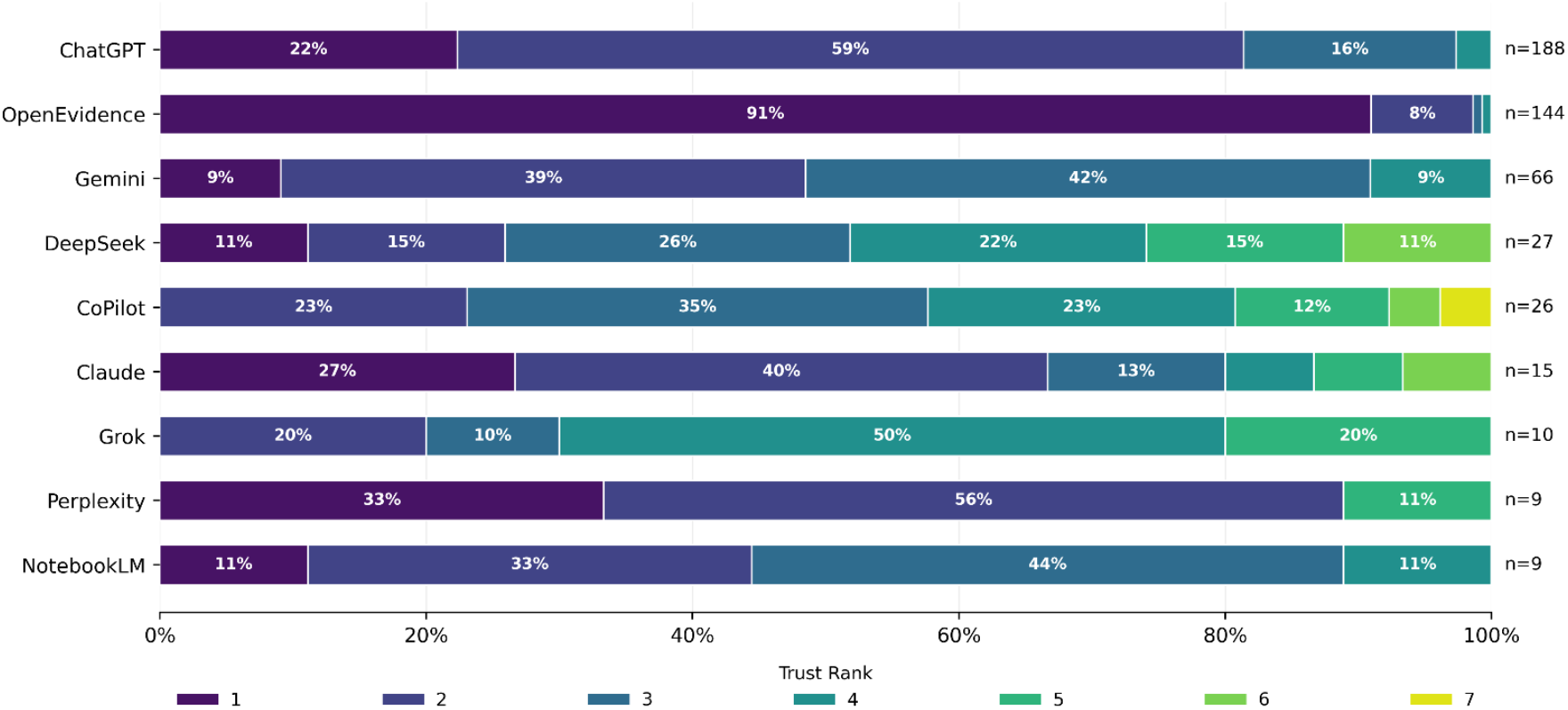
Frequency of highest trust ratings across used large language models. Respondents were asked to rank each large language model they had used in order of their trust. Displayed is the frequency of trust ratings. Only models with ≥5 respondent rankings are shown.

### Attitudes on Integration, Privacy, and Ethics

Most respondents agreed that formal LLM training should be integrated into curricula (180/266; 67.67%), but agreement distribution significantly differed by training stage (*p* = 0.043). No significant differences were observed between training stages for responses to other statements related to medical education integration. The majority of students agreed that lack of access to a subscription-based LLM creates barriers (141/266; 53.01%). Only 21.43% of students (57/266) felt adequately educated on data privacy regulations applicable to AI tools and respondents frequently endorsed ethical concerns regarding LLM use in medical education (55.64%; 148/266) (Figure 4).

**Figure 4.**
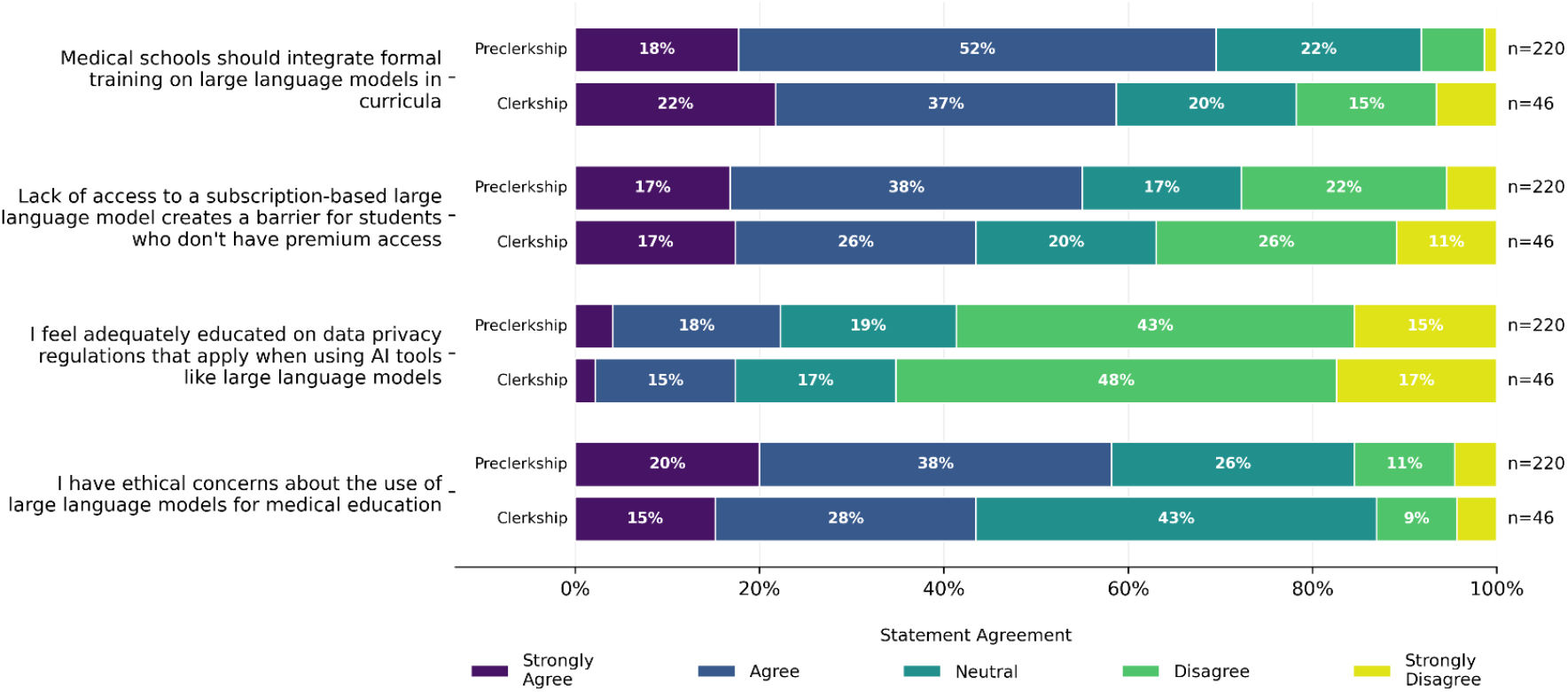
Agreement with statements related to curricular integration of large language models, stratified by training stage (preclerkship vs clerkship).

Most students (216/266; 81.20%) indicated that they believe LLMs will play a significant role in their medical career beyond medical school. Students indicated that they believe LLMs will have a positive impact on patient outcomes (180/266; 67.67%), but less frequently agreed with a positive impact on physician-patient relationships (111/266; 41.73%). Clerkship students were significantly more likely to indicate a positive impact on patient outcomes (*p* = 0.007). Other attitudinal questions related to clinical integration did not show significant differences between training stages. Students also frequently indicated ethical concerns about clinical integration (169/264; 64.02%) (Figure 5).

**Figure 5.**
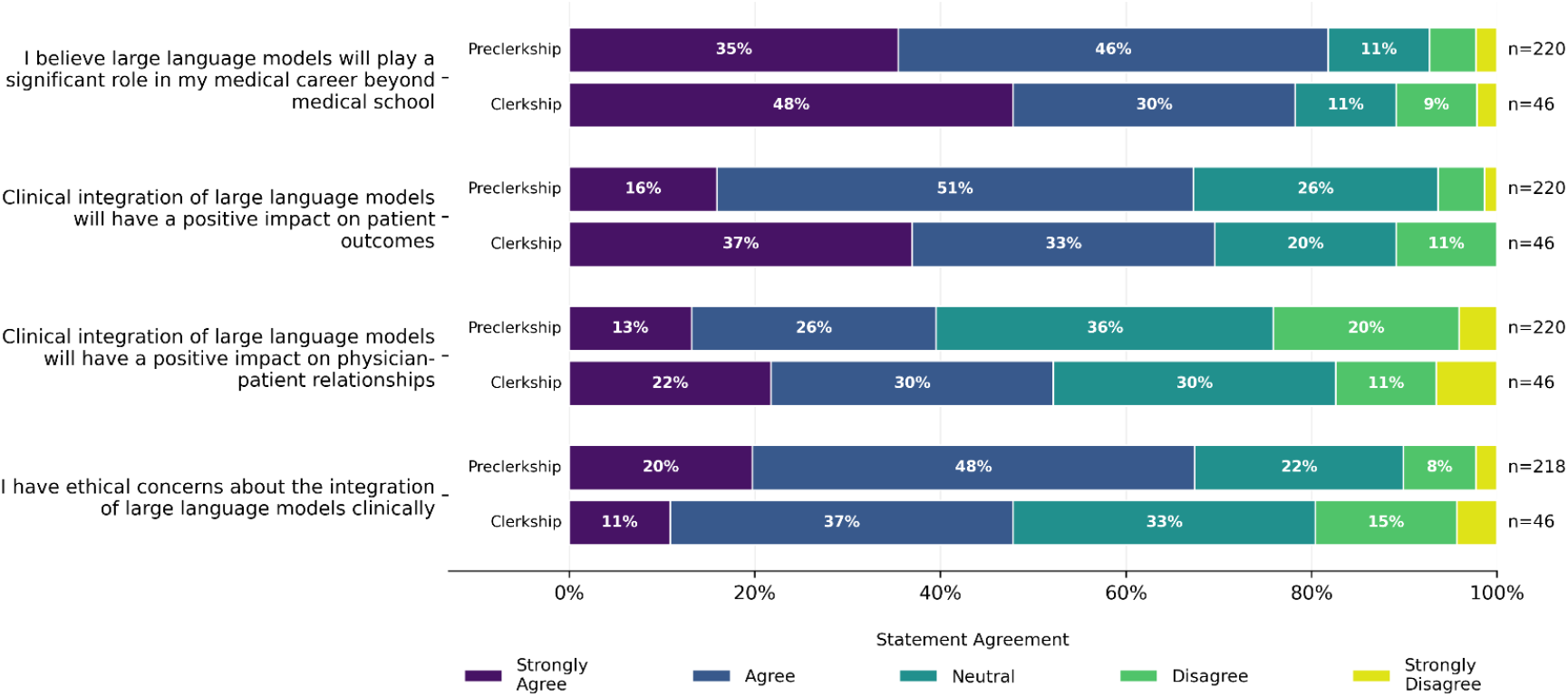
Agreement with statements related to clinical integration of large language models, stratified by training stage (preclerkship vs clerkship).

## Discussion

In this national survey of medical students in Canada, LLM use was nearly universal and deeply embedded in students’ learning and clinically oriented information seeking. Respondents reported frequent LLM use for coursework assistance and clinical questions, and most students perceived associated improvements in efficiency, learning, and academic performance. However, students also reported routinely encountering inaccurate outputs and described substantial gaps in formal instruction and privacy literacy, underscoring a widening mismatch between real-world adoption and structured educational governance.

Our observed prevalence of LLM use suggests that LLMs have shifted from novel adjuncts to baseline infrastructure in student workflows. This degree of penetration appears higher than prior survey data [20,21]. Internationally, surveys similarly describe widespread uptake among medical students, with common use cases including studying and writing support, alongside highlighted concerns of inaccuracy, bias, and privacy [20,21]. Our results extend this literature by providing contemporary Canadian data on which LLMs are used, usage patterns, comparisons by training stage, and student perceptions associated with their future practice as well as curricular and clinical integration.

Stage-of-training differences were also informative. Clerkship students reported greater use of OpenEvidence and were more likely to view LLMs as improving patient outcomes. OpenEvidence emerged as the most trusted tool among respondent trust rankings, across both stages of training. Preclerkship students, however, reported more frequent LLM use for coursework support and OSCE preparation, and were more likely to have been required to use an LLM during their schooling and to have encountered instructor-generated LLM content. While observed differences between preclerkship and clerkship students may reflect genuine variation in how learners engage with LLMs across training stages, they may also result from timing effects inherent to a cross-sectional survey conducted amid a rapidly evolving LLM landscape [3,22].

A central and concerning finding was the high prevalence of encountered inaccuracies. This aligns with well-described limitations of LLMs, including hallucinations and variable performance [1]. In a learning context, plausible-sounding but incorrect explanations can reinforce misconceptions and may be especially difficult for novice learners to detect. However, most respondents reported at least moderate trust in LLM outputs, which suggests a precarious relationship. The tension between known unreliability and perceived utility strengthens the argument for explicitly teaching information verification behaviours.

Despite widespread adoption, only a small minority of respondents reported receiving formal instruction on LLM use. This “training gap” mirrors concerns in the medical education literature that learner adoption is outpacing both policy and educator preparedness [23]. The gap may carry downstream consequences beyond academic integrity. Students reported lack of literacy regarding data privacy, a concern that gains greater saliency as LLM use extends into clinically adjacent settings, which could tempt inclusion of patient information in prompts.

Respondents had generally positive views of LLM utility in both medical education and clinical environments. Students commonly expressed positive perceptions of LLM implications on patient outcomes and physician-patient relationships. Students also largely indicated an expectation that LLMs will play a significant role in their future careers. Accordingly, integration of curricular guidance on LLM use can be framed as professional development, rather than merely a technical skills module.

This study has limitations. Participation was voluntary and disseminated through broad channels, which prevented calculation of a response rate and raised the possibility of selection bias. The sample overrepresented preclerkship respondents and included students from 10 of 18 Canadian medical schools; French-language programs were not represented. All measures were self-reported and may be influenced by recall or social desirability effects. In addition, model identification may be imperfect given the evolving branding and integration of tools. Finally, multiple exploratory comparisons were conducted between training stages; statistically significant differences should be interpreted cautiously and considered hypothesis-generating.

Future work should connect specific patterns of LLM use to objective outcomes, including knowledge acquisition, clinical reasoning performance, documentation quality, and downstream clinical decision-making. Mixed-methods research could clarify how learners verify outputs in real time, what prompts or workflows reduce error exposure, and which curricular interventions most improve safe use and privacy-protective behaviour. As LLM capabilities continue to expand into clinical documentation and workflow support, ongoing evaluation will be needed to ensure that medical education prepares learners to use these tools responsibly while preserving clinical reasoning and accountability. Given the near-ubiquitous and frequent LLM use observed here, studies should also explicitly evaluate potential impacts on trainee development—including deskilling, mis-skilling, and never-skilling, with particular attention to never-skilling risks when core competencies are offloaded before they may be consolidated [15-17].

## Conclusion

LLM use was nearly ubiquitous among surveyed medical students in Canada, and was frequently applied to coursework and clinically oriented questions. Students expressed benefits for learning, efficiency, and clinical outcomes, yet also reported frequent exposure to inaccurate outputs, low confidence in their data privacy literacy, and limited formal instruction on responsible use. These findings support the development of structured curricular training and institutional guidance addressing appropriate use, information verification, privacy, and ethics.

## Contributions

Conceptualization: AAB, RCR, MP, JKH; Data Curation: AAB; Formal Analysis: AAB; Investigation: AAB, RCR, KL, MP, ZK, ACM, AYH, MZ, OEF, AAK, PD, AH, KS; Methodology: AAB, RCR, MP, JKH; Project Administration: AAB, RCR, KL, MP, ZK, ACM, AYH, MZ, OEF, AAK, PD, AH, KS, MP, JKH; Supervision: MP, JKH; Visualization: AAB; Writing – Original Draft: AAB, RCR, KL, MP, ZK, ACM, AYH, MZ, OEF, AAK, PD, AH, KS; Writing – Review & Editing: AAB, RCR, KL, MP, ZK, ACM, AYH, MZ, OEF, AAK, PD, AH, KS, MP, JKH

## Data Availability

Individual participant data cannot be made available.

