## Supplementary Appendix 1 for "Medical Students’ Use of Large Language Models: A National Survey"

### Default Question Block

What stage of medical training are you currently in?

- ○ Preclerkship
- ○ Clerkship

School of Study

[ ▾ ]

Please select any of the following large language models you have used:

- ☐ ChatGPT
- ☐ Claude
- ☐ CoPilot
- ☐ DeepSeek
- ☐ Gemini
- ☐ Grok
- ☐ Llama
- ☐ Mistral

☐ OpenEvidence

☐ [ ] Other (please specify)

#### How comfortable do you feel using large language models?

| Extremely uncomfortable | Somewhat uncomfortable | Neutral | Somewhat comfortable | Extremely comfortable |
|---|---|---|---|---|
| ○ | ○ | ○ | ○ | ○ |

#### Have you ever been required to use a large language model for any part of your medical schooling?

○ Yes

○ No

#### What was the required use of a large language model in your medical schooling?

[ ]

Have you ever encountered large language model-generated content from instructors in your schooling?

- Yes
- No
- Unsure

Do you have a paid subscription to a large language model? If so, to what model(s)?

- [ ] Yes (please specify)
- No

For each of the following use cases, please indicate how often you use large language models to assist with each task:

| | Never | Rarely | Monthly | Weekly | Daily |
|---|---|---|---|---|---|
| Coursework assistance | ○ | ○ | ○ | ○ | ○ |
| Writing assistance | ○ | ○ | ○ | ○ | ○ |
| Clinical questions | ○ | ○ | ○ | ○ | ○ |
| Research | ○ | ○ | ○ | ○ | ○ |

| | Never | Rarely | Monthly | Weekly | Daily |
|---|---|---|---|---|---|
| Written examination preparation | ○ | ○ | ○ | ○ | ○ |
| OSCE preparation | ○ | ○ | ○ | ○ | ○ |
| Other (please specify) [ ] | ○ | ○ | ○ | ○ | ○ |

#### Have large language models impacted:

| | Negatively | No Impact | Positively |
|---|---|---|---|
| Your learning | ○ | ○ | ○ |
| Your academic performance | ○ | ○ | ○ |
| Your efficiency | ○ | ○ | ○ |

#### How do large language models compare to traditional learning resources (e.g., textbooks, modules) for learning?

| Much worse | Somewhat worse | About the same | Somewhat better | Much better |
|---|---|---|---|---|
| ○ | ○ | ○ | ○ | ○ |

How much do you trust the information provided by large language models?

Not at all | A little | Somewhat | Mostly | Completely

Rank the following large language models by your level of trust:

» ChatGPT

» Claude

» CoPilot

» DeepSeek

» Gemini

» Grok

» Llama

» Mistral

» OpenEvidence

» Other (please specify)

Have you encountered inaccurate information provided by a large language model?

- Yes
- No
- Unsure

If you have encountered inaccurate information, how often?

| Every prompt | Every few prompts | Once in a while | Rarely | Never |
|---|---|---|---|---|
| ○ | ○ | ○ | ○ | ○ |

Have you received any formal training or instruction on how to use large language models?

- Yes
- No

If you have received formal training or instruction on how to use large language models, from where?

- [ ] Medical school
- [ ] Work
- [ ] Online course
- [ ] Other (please specify)

Rate your agreement with the following statements:

| | Strongly disagree | Disagree | Neutral | Agree | Strongly agree |
|---|---|---|---|---|---|
| Medical schools should integrate formal training on large language models in curricula | ○ | ○ | ○ | ○ | ○ |
| Lack of access to a subscription-based large language model creates a barrier for students who don't have premium access | ○ | ○ | ○ | ○ | ○ |
| I feel adequately educated on data privacy regulations that apply when using AI tools like large language models | ○ | ○ | ○ | ○ | ○ |

| | Strongly disagree | Disagree | Neutral | Agree | Strongly agree |
|---|---|---|---|---|---|
| I have ethical concerns about the use of large language models for medical education | ○ | ○ | ○ | ○ | ○ |

#### Rate your agreement with the following statements:

| | Strongly disagree | Disagree | Neutral | Agree | Strongly agree |
|---|---|---|---|---|---|
| I believe large language models will play a significant role in my medical career beyond medical school | ○ | ○ | ○ | ○ | ○ |
| Clinical integration of large language models will have a positive impact on patient outcomes | ○ | ○ | ○ | ○ | ○ |
| Clinical integration of large language models will have a positive impact on physician-patient relationships | ○ | ○ | ○ | ○ | ○ |

| | Strongly disagree | Disagree | Neutral | Agree | Strongly agree |
|---|---|---|---|---|---|
| I have ethical concerns about the integration of large language models clinically | ○ | ○ | ○ | ○ | ○ |

#### Anything else you would like to share?

Powered by Qualtrics
